# Synaptic Vesicle Cycling Disorders: Cross-Sectional Phenotyping Study of a Gene Functional Network

**DOI:** 10.1101/2024.10.17.24315676

**Authors:** Josefine Eck, Tess Smith, Anna Kolesnik, Reem Al-jawahiri, Kate Baker

**Affiliations:** MRC Cognition and Brain Sciences Unit, University of Cambridge; John van Geest Centre for Brain Repair, University of Cambridge; Department of Neuroscience, Northwestern University Feinberg School of Medicine; GOS Institute of Child Health, University College London; Department of Medical Genetics, University of Cambridge; Department of Pathology, University of Cambridge

**Keywords:** Neurodevelopmental Disorders, Synaptic Vesicle Cycling, Presynaptic, Genetic, Phenotype

## Abstract

**Background and objectives:** Genes associated with neurodevelopmental disorders (NDDs) can be grouped into networks according to their molecular and cellular functions. However, the links between gene functional networks and neurodevelopmental phenotypes are not well understood. Synaptic vesicle cycling (SVC) is one gene functional network in which rare, high penetrance variants are known to cause NDDs. SVC genes regulate neurotransmitter release and recycling, thus essential for synaptic transmission and plasticity. We investigated whether SVC disorders are associated with a different neurodevelopmental spectrum from other monogenic NDDs, and whether phenotypic variation within SVC disorders can be predicted.

**Methods:** We included 199 children, young people, and adults with a genetic NDD diagnosis (n=109 with SVC disorders, n=90 with non-SVC disorders). Families were recruited via clinical genetics and neurology services, and support organisations. Parents or carers completed quantitative questionnaire measures previously validated in populations with NDDs. We implemented a PCA-derived K-prototype clustering analysis to assess associations between gene functional network and neurodevelopmental variation.

**Results:** Cohort-wide K-prototype clustering identified four phenotypic similarity clusters, each having membership from both SVC and non-SVC participants. The clusters differed in phenotype enrichments as follows: 1) behavioural difficulties and sleep problems, 2) less severe neurodevelopmental problems overall, 3) epilepsies, sleep problems, severe adaptive impairments and visual awareness difficulties, 4) sensory-motor problems. The SVC group was over-represented within cluster 3. Cluster memberships within the SVC group cannot be predicted by age, sex, variant type, gene or SVC sub-process.

**Discussion:** The K-prototype method can be used to describe multi-dimensional phenotypic structure within the heterogeneous genetic NDD population. We found that SVC functional network membership contributes to the likelihood of phenotype cluster memberships, but does not specify a distinct or homogeneous neurodevelopmental profile. Investigating convergent disease mechanisms arising from SVC dysfunction is central to understanding the observed neurodevelopmental spectrum, and may ultimately guide evidence-based prognostication and mechanism-informed management.

## Introduction

Neurodevelopmental disorders (NDDs) of known genetic origin are individually rare, but collectively common. NDDs caused by *de novo* variants have a collective incidence of 1 in 213-448 births^1^, with an ever-increasing number of monogenic NDDs discovered^2^. However, the links between NDD genetic aetiologies and phenotypic characteristics are not well understood^3^. One of the challenges faced is the rarity of each genetic diagnosis, given extreme genetic heterogeneity across the NDD population. Another challenge is phenotypic heterogeneity, whereby variations in the same gene can cause multiple different phenotypes of variable severity^4,5^. On the other hand, different monogenic NDDs often share overlapping characteristics^6,7^.

One approach that may help to improve our understanding of phenotypic and mechanistic architecture, is to explore groups of monogenic NDDs that converge on shared molecular and cellular functions. Such functional groups include postsynaptic signalling, chromatin regulation and cytoskeletal components^8^. These groups are expected to manifest similar characteristics (both homogeneity and variability) because of shared physiological and developmental mechanisms. This approach is known as gene functional network group (GFN) phenotyping, and is a starting point for uncovering shared multi-level mechanisms and ultimately therapeutic strategies that benefit a larger cohort of individuals than those affected by ultra-rare single gene disorders^9^. Previous studies have found evidence for associations between GFNs and neurodevelopmental phenotypes, applying quantitative comparative methods in individuals with rare monogenic NDDs^3,9,10^.

Synaptic vesicle cycling (SVC) is one such functional network in which rare, high penetrance variants are known to cause NDDs^11–14^. SVC genes encode proteins essential for synaptic transmission and plasticity, orchestrating release and re-cycling of neurotransmitter-filled vesicles (Figure 1)^15^. Vesicles are translocated to the presynaptic active zone where they attach to a dense protein network. To prevent excessive neurotransmitter release, synaptic vesicle fusion is actively inhibited at rest^15^. Action potentials induce the opening of voltage-dependent calcium channels, whereupon calcium influx triggers the fusion of docked and primed vesicles (i.e. exocytosis), facilitating post-synaptic receptor activation. Following exocytosis, vesicle-associated proteins and lipids are retrieved from the presynaptic membrane via various endocytosis modes (i.e. ultrafast, clathrin-mediated, activity-dependent bulk). Vesicles are then reformed, refilled and clustered into resting or recycling pools^8,14,16^.

**Figure 1.**
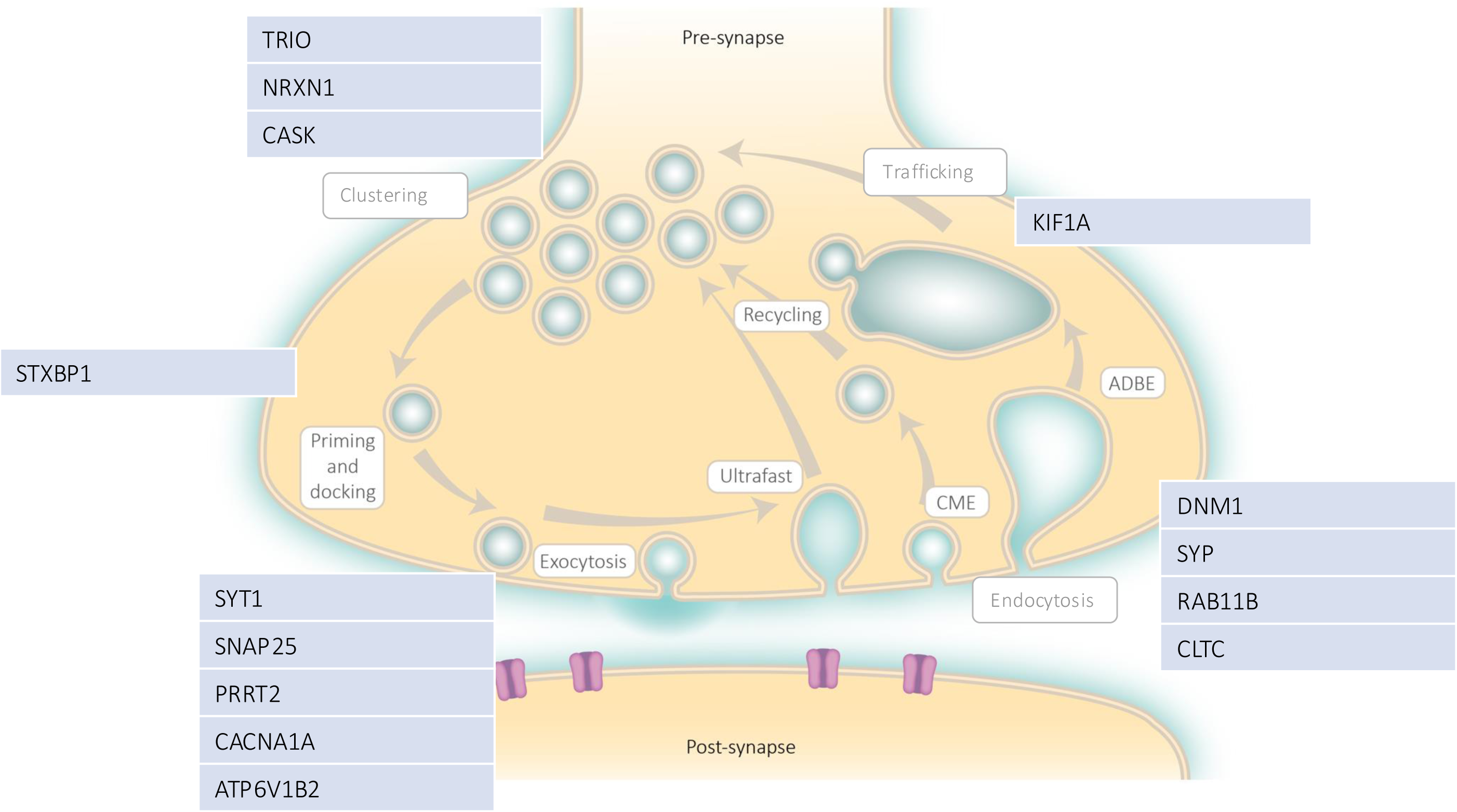
The life cycle of synaptic vesicles. The image was adapted and reproduced with permission from Bonnycastle et al., Journal of Neurochemistry, 2021 (John Wiley & Sons, Inc. © 2020 International Society for Neurochemistry). SVC genes represented in our cohort have been added to the figure. These genes are of clinical significance in neurodevelopment at different sub-processes of the tightly regulated synaptic vesicle cycle.

Clinical case series’ have indicated that NDD-associated SVC gene variants share a common set of neurodevelopmental manifestations^11^. These fall into four broad phenotype categories – developmental delay / intellectual disabilities (ID), movement disorders (MD), epilepsies and visual impairments^11,17^. The prevalence and severity of these manifestations vary between and within conditions, and are not an inevitable consequence of SVC disruption^11,14,16,18^. One such example is *STXBP1*-associated NDD, which was initially considered universally severe but is, in fact, more variable in age-of-onset, type and severity of presentations^4,5,14^. Similarly, the phenotypic spectra of *SYT1*-associated and *CASK*-associated NDDs includes a wider range of severities than initially reported^19,20^. These studies highlight the heterogeneity arising from variants in the same SVC gene, underlying the importance of careful phenotyping and cautious prognostication following diagnosis. The phenotypic characteristics of SVC disorders have not been systematically described in comparison to other monogenic NDDs. Therefore, it is not known whether characteristics are more similar in types or severities amongst the SVC group than expected for any monogenic NDDs.

It remains unclear how presynaptic dysfunction can lead to numerous phenotypes with wide-ranging severity. Potential factors that could mediate variability include variant type, gene expression topography and timing. For example, *VAMP2* missense variants have been associated with more severe phenotypes than frameshift variants^23^, whereas the reverse may be true for *CASK*, indicating the potential for contrasting impact of gain versus loss of function, as observed for channelopathies and post-synaptic receptors^21^. Specific SVC genes (or potentially specific variants) can have predominant impact on one or more SVC physiological sub-processes, exerting phenotypic influence. Systematic literature review indicated that SVC genes involved in fusion, endocytosis and trafficking were more frequently associated with visual dysfunction than priming and docking disorders - disorders of endocytosis (*CLTC, DNM1, RAB11A*) were always associated with visual impairment whereas visual symptoms were more variable in other conditions^11^. The presence of some clinical symptoms may influence the emergence or severity of other symptoms. Visual impairment is particularly common among SVC individuals with motor impairments^11,22^ - these two phenotypes may co-occur because they arise from similar neuroanatomical or physiological systems, or may reflect global severity, or may make independent contributions to cognitive development.

In summary, mapping the phenotypic landscape of SVC disorders is required to inform future investigation of SVC-related molecular functions, neuronal physiology and developmental cognitive systems. Here, we adopt a GFN phenotyping approach to test the proposal that SVC disorders share convergent neurodevelopmental characteristics^11^. Our primary objective is to determine whether SVC disorders are associated with a different neurodevelopmental spectrum (in type or severity) from other monogenic NDDs. Our secondary objective is to explore whether neurodevelopmental variation within SVC disorders is associated with predictive factors within the group. To meet these objectives, we apply a dimensional clustering approach to quantitative phenotyping measures, identifying clusters of phenotypic similarity across the NDD cohort, and then assessing associations between potential predictive factors and cluster membership.

## Methods

### Participants and recruitment

We present a cross-sectional case-control study of 199 children, young people and adults who had been clinically diagnosed as having a neurodevelopmental disorder (NDD). A further inclusion criterion was a genetic diagnosis obtained via clinical genetics services judged to be definitely or likely causal or contributory to participants’ NDD. We included pathogenic variants, likely pathogenic variants and variants of uncertain significance based on diagnostic-grade ‘green’ genes on PanelApp England, an evidence-based open-access online database used to facilitate diagnostic variant filtering^23^. Participating families received publicity about the study from regional clinical services or via support organisations, and self-referred via the project website. Participants were located internationally; all data were collected by the U.K.-based study team.

Participants were allocated to GFNs based on the molecular and biochemical function of their diagnostic genetic variant. In this study, we allocated genetic disorders converging on presynaptic dysfunction to the ‘SVC group’ (*Supplementary Table 1*), based on strong evidence for direct involvement in vesicle cycling, and pathophysiology relating to SVC disruption^11,12,24,25^. All other monogenic NDDs were allocated into the ‘non-SVC group’ (*Supplementary Table 2*).

### Questionnaire measures

Phenotyping assessments were selected to be appropriate for populations with NDDs and the age range of participants, and practical for remote completion. Parent-report questionnaires were completed online or by post or online interview according to parent preference. Clinical information was collated from medical documentation and the medical history questionnaire (MHQ) which covered perinatal history, infant and child health, current health and developmental milestones. Global and domain-specific adaptive functioning was measured by the Vineland Adaptive Behaviour Scales, Second or Third Edition (VABS)^26,27^. The Social Responsiveness Scale, Second Edition (SRS-2)^28^ was included as a measure of social communication and interaction, restricted interests and repetitive behaviour. VABS and SRS raw scores were scaled according to published normative data. The Developmental Behaviour Checklist, Second Edition (DBC-2)^29^ was employed as a measure of behavioural and emotional difficulties designed for individuals with NDDs. Raw scores were converted to T-scores and stratified by ID severity derived from the VABS composite score. Visual impairment and its functional impact were measured using the Cerebral Visual Impairment scale (CVI)^30^.

### Statistical analyses

We first summarised demographic variables, MHQ-derived descriptive variables and standardised scores from the VABS, DBC, SRS, and CVI for both groups. Using Fisher’s Exact Test, we explored whether the two groups differed significantly in the frequency of MHQ-reported clinical phenotypes.

To investigate quantitative adaptive, social, behavioural and visual characteristics that might differentiate SVC disorders from other NDDs, we applied Principal Components Analysis (PCA; SPSS v29.0.2.0) to reduce the large number of variables into principal components that best represent the original data^31^. Prior to PCA, we standardised the dataset by converting normed subscale scores to a percentage of the maximum score across the cohort. We inversed VABS scores to aid interpretation. Missing data were imputed for continuous variables using the MICE package in R. The Kaiser-Meyer Olkin measure assessed sampling adequacy and Bartlett’s Test of Sphericity determined the appropriateness of PCA. The criteria for retaining components were (1) the Eigenvalue-one criterion, (2) ≥ 5% total variance explained, (3) a pronounced elbow in the scree-plot and (4) the rotated component matrix values ≥.5 as significant contributors to variance^32–34^. To formally evaluate the predictive capability of the PCA solution, 5-fold cross-validation (CV) was carried out using the scikit-learn package in Python. In k-fold CV, k-1 folds are used for model building, while the remaining fold is designated for validating the model. Previous studies have adopted k-fold CV to select optimal component number to the lowest root mean squared error^32^, generating more stable predictions and removing redundant data. To investigate differences in PCA factor component scores between SVC and non-SVC groups, normality for each component was tested, followed by pairwise comparisons.

We then determined whether there are phenotypic similarity clusters of individuals with SVC and non-SVC disorders, and explored predictive factors influencing cluster memberships. To implement this, we combined extracted PCA factor scores and clinical data, using the K-prototype algorithm. K-prototype clustering incorporates K-modes and K-means clustering to effectively cluster datasets containing a combination of categorical and continuous variables^35^. We included all five PCA factor scores (namely social-emotional, adaptive functioning, behavioural, sensory-motor, and visual awareness), and clinical data assessed across the cohort (namely feeding, tone, sensory, self-injury, epilepsy and sleep) as K-prototype variables. The algorithm starts by randomly selected cluster centroids, then iteratively assigning each data point to the nearest cluster centroid using a chosen distance metric, updating the centroids until convergence. K-prototype clustering was carried out using the kmodes package in Python with Huang initialisation method^36^. We implemented the Euclidean distance as a parameter for continuous data and the matching dissimilarity measure for categorical data. We employed the elbow method to select the optimal number of clusters^35,37^. The curve’s inflexion point (elbow) serves as a reliable indicator for identifying cluster divisions, optimising the cost function thereby minimising within-groups sum of squared errors.

To inspect phenotypic differences between each identified cluster, categorical and continuous variables were analysed using chi-square analyses and Analysis of Variance (ANOVA), respectively. We tested whether cluster membership within the combined cohort could be predicted by demographic factors (age, biological sex), GFN or variant type. We then explored whether neurodevelopmental variation within SVC disorders is associated with specific predictive factors by removing all non-SVC participants and re-analysing the cohort-wide cluster solution. Potential predictors of cluster membership were as above, with the addition of MDs and SVC sub-processes.

### Standard protocol approvals, registrations, and patient consents

Phenotyping data were collected within the ‘Phenotypes in Intellectual Disability’ research study at the University of Cambridge, which received ethical approval from the Cambridge Central Research Ethics Committee (REC ref: IRAS 83633). Written informed consent was provided by a parent or consultee for all participants.

### Data availability

The data that support the findings of the present study are not openly available due to reasons of sensitivity and are available to other ethically approved research projects upon request from the corresponding author. Data are located in controlled access data storage at the MRC Cognition and Brain Sciences Unit, University of Cambridge.

## Results

### Descriptive results

*Supplementary Table 3* provides demographic and questionnaire data for the cohort. The SVC group comprised 109 individuals (65 female, 44 male), aged 2.67 to 32.33 years (M= 10.63; SD= 6.43). The non-SVC group comprised 90 individuals (49 female, 41 male), aged 2.69 to 26.60 years (M= 12.92; SD= 5.65). *Supplementary Tables 4* and *5* detail clinical phenotypes within the SVC and non-SVC groups. A high proportion of both groups have gastrointestinal symptoms, motor delay and speech delay. There were no significant differences between groups in the frequency of eye physiology problems, tone abnormalities, sleep disturbances, feeding difficulties, prenatal concerns or / perinatal problems (*Supplementary Table 6*). Seizures were reported in half of participants in the SVC disorders group, but only 28% of participants in the non-SVC disorders group (*p= .003, Cramer’s V= .218*). Self-injurious behaviour (including finger biting or chewing, head banging and skin picking) was reported in 59% of SVC individuals and 40% of non-SVC individuals (*p= .034, Cramer’s V= .191*). Individuals in the SVC group were significantly more likely to have parent-reported movement disorders (*p= .025, Cramer’s V= .227*). Across all behavioural questionnaire measures, the spread of data suggests substantial variability within both groups.

### Questionnaire analysis – PCA and pairwise comparisons

Missing data were imputed which accounted for 15.3% of total missingness (*Supplementary Figure 1*). We carried out a varimax-rotated unified PCA using imputed subscale measures. Bartlett’s test of sphericity reached statistical significance (*p* = <.001), indicating that the dataset was likely factorizable. There was a pronounced elbow in the scree plot after five factors, demonstrating that there was little explanatory benefit (i.e. principal components with eigenvalues < 1) in extracting more than five factors (*Supplementary Figure 2*). The PCA generated a robust result (Kaiser-Meyer-Olkin= .809) with a five-factor rotated solution that accounted for 70.68% of total variance (*F1= 29.32%, F2= 16.14%, F3= 10.66%, F4= 7.78%, F5= 6.80%*). Figure 2 shows the loadings of each questionnaire subscale to each factor component (*Supplementary Tables 7 and 8*). 5-fold CV confirmed that the 5-factor rotated PCA solution provides a reliable estimate of model performance. The first component (explaining largest amount of variance) comprised all SRS subscales and the DBC social relating subscale. The second component comprised VABS subscales. All DBC subscales except social relating loaded onto the third component. The fourth and fifth principal components were composed of CVI subscales. These included items related to sensory-motor and visual awareness, respectively.

**Figure 2.**
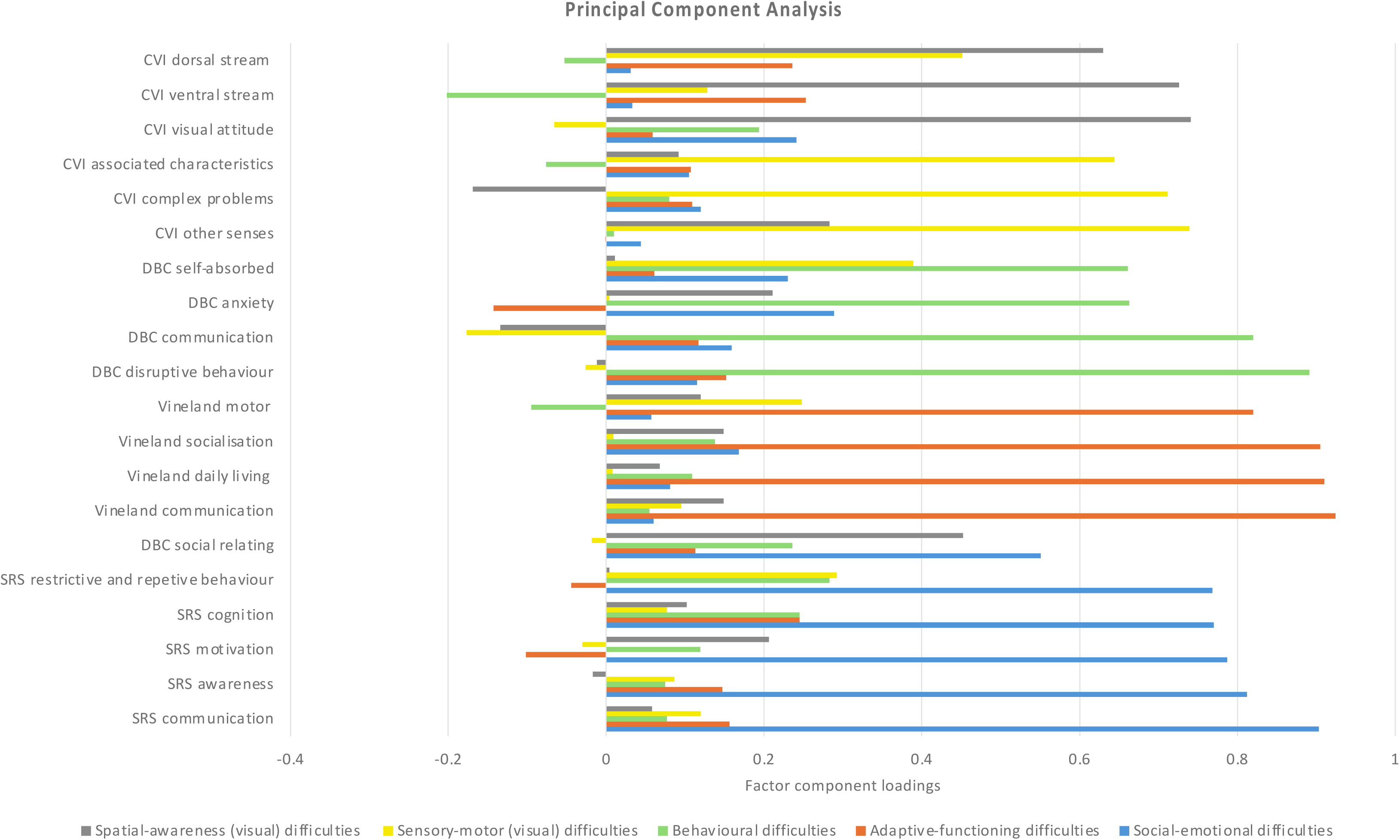
Contribution of questionnaire subscale measures to each factor of the unified PCA. Bars represent the factor loadings of each subscale measure onto each extracted component. Factor loadings represent the weighting of each subscale on each factor. Different colours represent different factor components, starting from the bottom with the first principal component explaining the largest possible amount of variance in the data. Significant contributors of variance (set at ≥0.5) are indicated as values extending outside the grey box.

Pairwise comparisons were carried out to inspect whether PCA factor scores differed between groups. Results revealed statistically significant differences in adaptive functioning (*U= 3322, Z= −3.915, p= <.001*), indicating, on average, more severe adaptive impairments in the SVC group (*Mdn=.34, n= 109, 95% CI= −1.42, 2.82*) than in the non-SVC group (*Mdn= −.38, n= 90, 95% CI= −1.52, 2.04*). We also found significant differences in problems related to visual awareness (*t(196)= −2.15, p= .033, d= .99*), with more severe impairments, on average, in the SVC group (*M= .134, n= 109, SD= 1.09, 95% CI= −.07, .34*) compared to the non-SVC group (*M= −.162, n= 90, SD= .85, 95% CI= −.34, .02*). We did not find any statistically significant differences between groups in social-emotional (*U= 4353, Z= - 1.365, p= .172*), behavioural (*t(197)= 1.069, p= .287, d= .99*) or sensory-motor difficulties (*U= 4641, Z= 8736, p= .514*).

### Cluster analysis – Cohort-wide phenotypic similarity clusters

Cluster analysis was performed using the K-prototype algorithm, combining PCA factor components with clinical data. Due to missing clinical data for 20 individuals, the final analysis included 179 individuals (*SVC group= 100; non-SVC group= 79*). K-prototype clustering classified the cohort into 4 clusters. Further subdivision yielded little additional explanatory power, indicating that a 4-cluster solution was best supported by the data. Table 1 and Figure 3 illustrate the relative clinical and behavioural characteristics of participants within the 4-cluster solution. Cluster 1 features higher prevalence or severity of behavioural problems, sleep problems and visual awareness problems.

**Figure 3.**
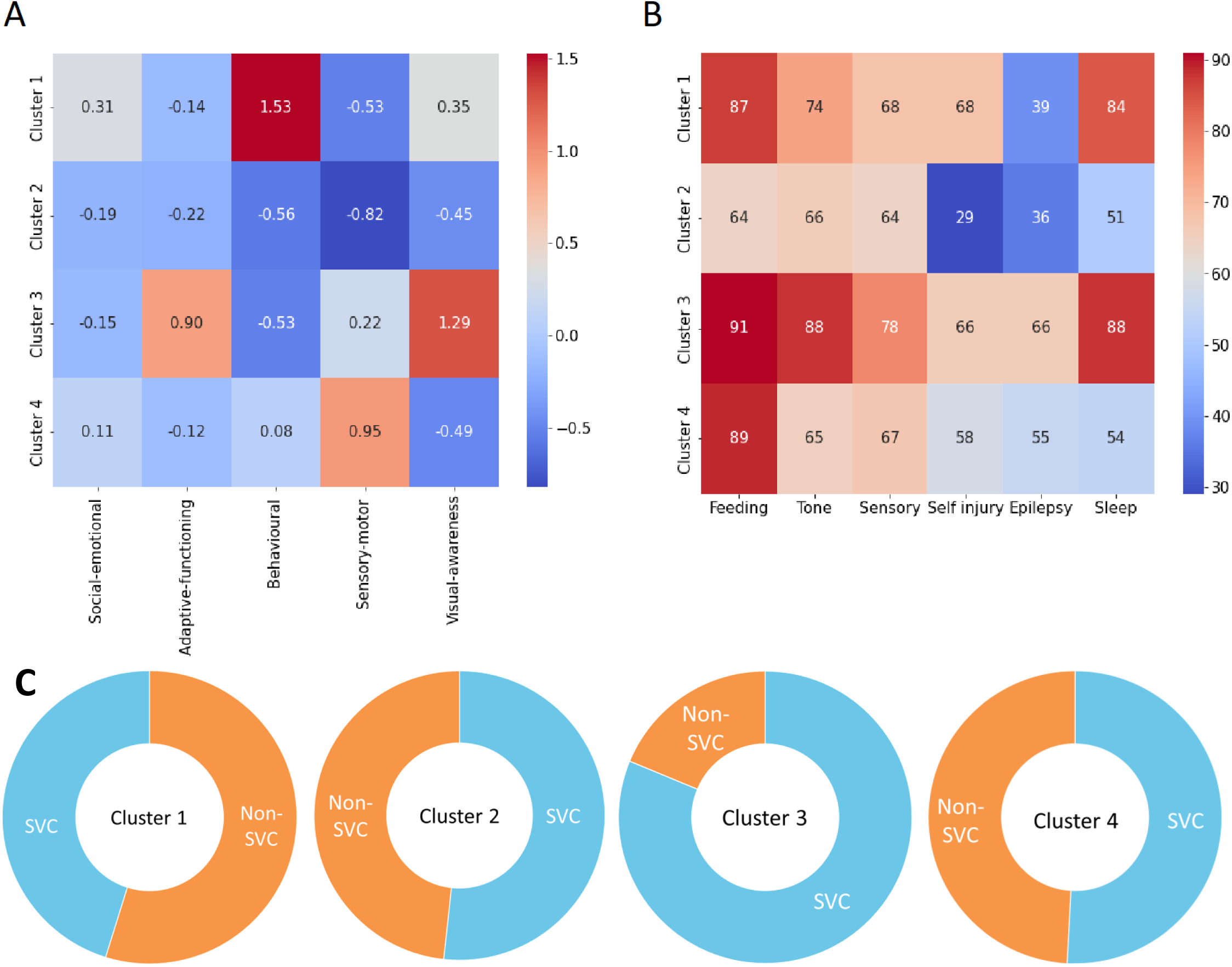
K-prototype cluster characteristics (combined cohort) (A) Neurodevelopmental dimensions derived from individual factor component scores of the unified PCA for the cohort-wide 4-cluster solution. Warmer colours indicate more difficulties. (B) Frequency of clinical symptoms derived from the MHQ and reported as percentages within each cluster. Warmer colours indicate a higher frequency of reported symptoms. (C) Gene functional network group membership, proportions within each cluster.

**Table 1.** Demographic information for the K-prototype 4-cluster solution.

| Cluster | SVC<br>n (%) | non-SVC<br>n (%) | Total<br>n | Age |  |  | Biological sex |  | Variant type |  |
| --- | --- | --- | --- | --- | --- | --- | --- | --- | --- | --- |
|  |  |  |  | Mean | SD | Range | Female | Male | PTV | Missense |
| Cohort-wide |  |  |  |  |  |  |  |  |  |  |
| 1 | 14(45) | 17(54) | 31 | 14.61 | 6.81 | 4-32 | 14 | 17 | 5 | 11 |
| 2 | 31(53) | 28(47) | 59 | 11.49 | 5.83 | 4-26 | 38 | 21 | 21 | 18 |
| 3 | 26(81) | 6(19) | 32 | 11.00 | 6.26 | 3-27 | 19 | 13 | 8 | 17 |
| 4 | 29(51) | 28(49) | 57 | 10.14 | 5.90 | 3-29 | 34 | 23 | 14 | 25 |
| SVC-only |  |  |  |  |  |  |  |  |  |  |
| 1 | 14 | - | - | 11.86 | 7.12 | 4-32 | 7 | 7 | 1 | 7 |
| 2 | 31 | - | - | 10.10 | 6.12 | 4-26 | 21 | 10 | 12 | 12 |
| 3 | 26 | - | - | 10.88 | 6.65 | 3-27 | 16 | 10 | 8 | 14 |
| 4 | 29 | - | - | 9.93 | 6.76 | 3-29 | 17 | 12 | 11 | 15 |

Participants in cluster 2 had significantly fewer problems in behavioural and visual functioning, and showing significantly fewer problems related to feeding, self-injury, and sleep. Cluster 3 is characterised by more severe adaptive functioning and visual awareness problems, sleep disturbances and epilepsy. Cluster 4 was characterised by elevated sensory-motor problems and behavioural problems. Clusters did not differ in social-emotional factor scores.

We conducted statistical analyses to determine which demographic and genetic factors influence cluster membership (*Supplementary Table 9*). We did not find any statistically significant differences between the clusters in biological sex distributions. Cluster 1 showed significant increase in age, driven by older participants (Tukey’s HSD; *95% CI [12.11, 17.11]*) compared to younger participants in cluster 4 (*95% CI [8.58, 11.71]*). Proportion of individuals with different variant types (missense versus protein-truncating) did not differ across clusters. The distribution of genes across each cluster is illustrated in *Supplementary Figure 3*. Considering GFN membership, there was an overall difference between SVC and non-SVC disorders in distribution across the four clusters (*x*^2^ *= 10.6, df = 3, p = .014, Cramer’s V = .244*). Cluster 3 had a significant overrepresentation of SVC participants with 81% of its members being from the SVC group compared to 19% from the non-SVC group. In contrast, the remaining clusters displayed a relatively even distribution of SVC and non-SVC individuals.

### Within-SVC-group phenotypic variability

Given the within-SVC group phenotypic variability, we explored the factors that predict neurodevelopmental and clinical variation (i.e. cluster membership) within this group, using the cohort-wide cluster solution (Table 1; Figure 4). Cluster 1 (14%) was characterised by significantly more behavioural problems. Individuals in cluster 2 (31%) reported fewer problems overall, with relatively reduced behavioural problems, sensory-motor functioning, and visual awareness. Cluster 2 additionally had the lowest proportion of feeding difficulties, self-injurious behaviour, and sleep problems. Cluster 3 (26%) was associated with significantly poorer adaptive functioning, visual awareness, feeding difficulties, and sleep problems. Cluster 4 (29%) reported the highest proportion of behavioural problems, sensory-motor problems and self-injury. There were no statistically significant differences between clusters in social-emotional functioning, tone abnormalities, sensory problems, epilepsies, or movement disorders. We assessed potential demographic or genotypic predictors of within-SVC cluster membership. Cluster membership did not differ significantly by age, biological sex, variant type or SVC sub-process. The genes occupying each cluster were relatively evenly distributed, including spread of the more prevalent SVC genetic diagnoses across clusters (*Supplementary Figure 4*).

**Figure 4.**
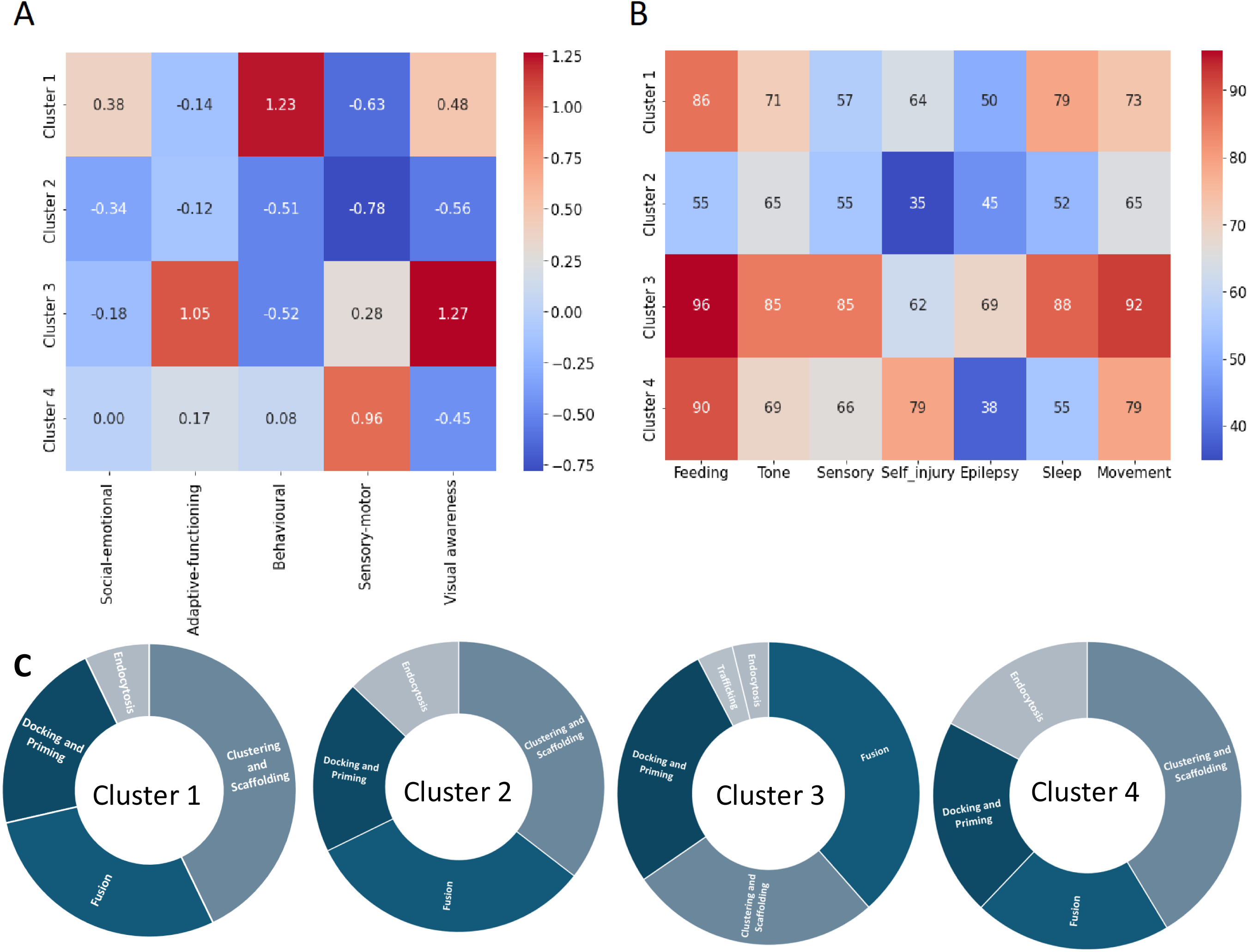
K-prototype cluster characteristics (SVC disorders group). (A) Neurodevelopmental dimensions derived from individual factor component scores of the unified PCA for the cohort-wide cluster solution (SVC disorder participants only). Warmer colours indicate more difficulties. (B) Frequency of clinical symptoms derived from the MHQ and reported as percentages within each cluster (SVC disorder participants only). Movement disorders are displayed on the heatmap but were not included as a prototype variable in the cluster solution due to a high percentage of missing data for the non-SVC group. (C) Genetic diagnoses grouped by predominant SVC sub-process, proportions within each cluster.

## Discussion

Neurodevelopmental disorders (NDDs) have traditionally been classified either by purely behavioural criteria (for example DSM diagnoses of ID, ADHD, autism etc.) or by aetiology, which may be genetic (for example trisomy 21, Fragile X syndrome, Rett Syndrome etc.), non-genetic (for example prematurity, Fetal Alcohol Syndrome) or multifactorial. However, neither behavioural nor aetiological classifications can fully explain the complexity of NDDs, encompassing within-diagnosis diversities and cross-diagnosis overlaps. These limitations in current concepts and knowledge are problematic given increasing availability and yield of diagnostic genetic testing, with growing numbers of individuals receiving a rare genetic NDD diagnosis and growing expectations that genetic diagnosis will inform prognosis and management. Ultimately, integration of aetiology with clinical characteristics to understand developmental systems contributing to each individual’s dynamic strengths and difficulties will be helpful.

One route toward defining these developmental systems may be to consider the phenotypic landscape of genetic NDDs converging on shared molecular and cellular mechanisms i.e. gene functional networks (GFN). Toward this goal, we carried out a data-driven dimensional phenotyping study of NDDs arising from rare variants in genes involved in presynaptic vesicle cycling (SVC disorders) and a mixed genetic NDD comparison group. We aimed to describe the neurodevelopmental spectrum of SVC disorders, to assess whether there is greater similarity than expected for the broader genetic NDD population, and to explore contributors to variability within the SVC disorders group.

Systematic parent-reported data indicated that, whilst no symptom was specific to or universal within the SVC disorders group, individuals with SVC disorders were more likely to have movement disorders (MDs), epilepsies and self-injurious behaviours. These results provide quantified evidence consistent with previous clinical studies ^5,11,12,19,38,39^. Caution is always warranted with regard to biases in clinical and research ascertainment, however the comparison non-SVC group was recruited via the same pathways for the same eligibility criteria and assessed via the same methods, with matched demographics and a similar distribution of genetic diagnosis numbers. Hence these findings suggest that dysregulation of SVC physiology influences developmental systems contributing to motor control and seizure susceptibility, differentially from other genetic NDDs.

MDs encompass a broad range of motor symptoms and reflect damage to or abnormal development of the basal ganglia or cerebellum and associated neural networks. Previous neuroimaging studies of SVC disorders have not uncovered consistent structural brain abnormalities^11,17^, however cerebellar atrophy has been observed in association with calcium channelopathies^40^ and distributed subcortical-cortical network abnormalities have been observed in association with *PRRT2* variants^41^. GFN-wide quantitative structural and functional neuroimaging studies may be helpful in delineating the origins of diverse MD symptoms amongst this group. Self-directed behaviours can have complex origins in neurodevelopmental disorders reflecting motor stereotypies and involuntary disinhibition, sensory stimulation or expression of frustrations. These symptoms warrant further investigation within SVC disorders because they are common, disruptive, distressing and difficult to ameliorate.

There is a very broad spectrum of epilepsies within the SVC disorders GFN and within specific genetic diagnoses, varying in seizure types, frequency, age of onset and treatment responses^5,11,42–45^. It is not yet known whether there is a shared set of mechanisms contributing to seizure susceptibility or epilepsy evolution within the SVC disorders group, distinctive from other synaptopathies or developmental encephalopathies more broadly. Previous studies of genotype-phenotype correlations for epilepsy within SVC disorders have found inconsistent effects of variant type. Truncating variants in *DNM1*, *SNAP25* and calcium channel subunits have been associated with more severe epilepsy phenotypes than missense variants^46,47^, whilst this was not found to be the case for *VAMP2*^5^. This could indicate divergent mechanisms of epileptogenesis for different SVC disorders, or opposing variant effects converging on a common pathway of abnormal excitability and circuit development. Larger samples are required to resolve these questions.

Beyond these important neurological symptoms, the major determinants of quality of life for patients and families affected by NDDs are cognitive, communicative and social-emotional functions. We mapped these functions using quantitative parent-report questionnaire measures, to assess whether there is a single dimension of severity or multiple relatively-independent dimensions, which may be differentially affected. We applied a varimax-rotated PCA to neurodevelopmental measures which indicated that ∼70% of total variation across the cohort (SVC and non-SVC disorders combined) can be captured by five orthogonal dimensions namely social-emotional, adaptive functioning, behavioural, sensory-motor and visual awareness abilities. To ensure our models and predictions remained unbiased, we conducted a 5-fold CV procedure, which corroborated PCA results. Using PCA, we were able to minimise the influence of variation related to noise, reduce dimensionality, and extract factor component scores, thereby facilitating subsequent analysis and data interpretation^48^. These results suggest that multiple dimensions characterise the genetic NDD population, and there may be relative independence in how these dimensions are impacted. Adaptive functioning and visual awareness dimensions were on average more severely affected amongst members of the SVC group, whilst other dimensions were not different on average between groups. It is also noteworthy that sensory-motor impairments were not more prominent in the SVC group compared to the non-SVC group, suggestive of disproportionate visual processing problems in SVC disorders. Future studies of SVC-associated visual dysfunction deploying electrophysiological methods may be a particularly interesting avenue to explore further.

Beyond symptom prevalence and dimensional differences between groups, we wanted to generate a more comprehensive and holistic view of neurodevelopmental co-occurrence and variation across this genetic NDD population. To achieve this, we used K-prototype clustering blind to genetic diagnosis, which grouped participants based on similarities in neurodevelopmental and clinical profiles. This identified a 4-cluster solution, each cluster combining clinical and questionnaire variables to identify sub-populations with similar characteristics. Interestingly, these clusters did not simply reflect four gradations of global severity across all measures; neither was there a sub-population dominated by high scores on the SRS screening questionnaire for autism i.e. the cluster solution does not map onto traditional behavioural categories. Cluster membership was not influenced by variant type or biological sex, and individuals with SVC disorders and specific genetic diagnoses were distributed across all 4 clusters, indicating that there is no homogeneous phenotypic profile predicted by diagnosis of any SVC disorder. However, relative to the distribution of non-SVC group members across clusters, the SVC disorders group were more likely to be found in cluster 3. Aligning with our knowledge of individuals with SVC disorders^11,19^ and pairwise comparisons of study data, this cluster represents individuals with severe adaptive functioning and visual awareness problems. This result has some, though limited, clinical utility by emphasising both the variability of prognosis for SVC disorders and the higher likelihood of severe cognitive difficulties without higher likelihood of autistic or emotional-behavioural difficulties. For future neuroscientific enquiry, this result can focus attention on the relationships between SVC physiology, visual processing and cognitive development.

Given the observed cluster distribution of SVC disorder participants, we explored whether there were predictors of cluster membership within the group. Considering potential clinical predictors, there were no significant differences between the SVC cluster groups in tone abnormalities, epilepsies or movement disorders. We also found no effects of age, biological sex or variant type on cluster membership. A recent study has highlighted more severe adaptive functioning in *VAMP2* missense variants than in *VAMP2* protein-truncating variants^18^, potentially as a result of GoF or specific dominant-negative effects^17^. Contrastingly, these variant-specific effects were not observed for *STXBP1* ^4,5,49^ aligning with the current results. Furthermore, no single genes dominate each cluster, indicating the absence of gene-specific effects within the current study. Classification of the SVC disorders group according to SVC sub-processes thought to be maximally dependent on each gene was also not informative in this initial analysis, although sub-process groups were not balanced within the sample. One hypothesis for future testing is that individual SVC gene variants can have highly specific effects on presynaptic physiology, not predictable in a straightforward manner by gene or missense / PTV. Qualitative and quantitative differences in physiological impact may predict cognitive outcomes, as recently observed for *SYT1*-associated NDD^50^. Testing of this hypothesis requires objective and systematic analysis of variant-specific functional effects and correlation with human phenotyping to understand the variable impacts of SVC gene variants on cognitive development.

### Strengths, limitations, and future directions

A strength of this study is the detailed post-diagnostic assessment of neurodevelopmental characteristics using standardised questionnaires alongside clinical and medical summaries of a relatively large cohort of individuals with monogenic NDDs. Another strength is our statistical methodology. Although K-means is widely used for clustering, its simplicity limits its use in more complex clustering tasks. Here, we extended the K-means algorithm by employing a less restrictive alternative method that retains the key properties of an underlying iterative deterministic model. Given that many individuals with monogenic NDDs present with complex phenotypes, using K-prototype clustering enabled us to achieve more meaningful and interpretable clusters. However, there are several caveats. First, we relied on parent-reported questionnaires and interviews, limiting the sensitivity, specificity and scope of our data. For instance, parents or caregivers may not have been acquainted with the specific medical jargon within questionnaires, and therefore under-reported or over-reported the presence of clinical characteristics in their child. Whilst questionnaire methods are common in this field of research, we strongly advise future work to incorporate multi-informant reports, direct observations and neuropsychological assessments. Sample sizes remain small for each specific genetic diagnosis, and are not balanced across SVC genes, however this may never be possible in view of ultra-rarity hence the GFN approach aims toward generalisability. Additionally, our study population spanned a broad age range, and we present cross-sectional results. It is important for future research to include longitudinal studies to explore the developmental trajectories within and between SVC disorders and genetic NDDs more broadly. This approach would shift from a categorical to a continuous phenotyping perspective, recognising that phenotypes are shaped by complex, contingent and fluid mechanisms rather than representing static and discrete states.

## Data Availability

The data that support the findings of the present study are not openly available due to reasons of sensitivity and are available to other ethically approved research projects upon request from the corresponding author.

## Acknowledgements

The authors sincerely thank the study participants, their families, and carers for their dedication and time in contributing to this study. We also would like to acknowledge the numerous clinicians, laboratory scientists and research personnel who identified potential participants and publicised this study. We also acknowledge the British Paediatric Neurology Surveillance Unit for assistance with recruitment to the study. We are grateful to general practitioners, local paediatricians, neurology, and neurophysiology services for facilitating access to medical records. This project was supported by funding from UK Medical Research Council (MC_UU_00030/3), the Great Ormond Street Hospital Charity and NIHR Cambridge Biomedical Research Centre.

