## Supplementary Material for "Synaptic Vesicle Cycling Disorders: Cross-Sectional Phenotyping Study of a Gene Functional Network"

|  |  | Page |
| --- | --- | --- |
| Supplementary Table 1 | *Genetic diagnoses in the SVC group* | 2 |
| Supplementary Table 2 | *Genetic diagnoses in the non-SVC comparison group* | 3 |
| Supplementary Table 3 | *Demographic information and questionnaire descriptive data* | 4 |
| Supplementary Table 4 | *Clinical phenotypes summary across the SVC-disorder group* | 5 |
| Supplementary Table 5 | *Clinical phenotypes summary across the non-SVC disorders group* | 7 |
| Supplementary Table 6 | *Comparison of clinical characteristics between SVC and non-SVC groups* | 9 |
| Supplementary Figure 1 | *Data missingness prior to imputation for all neurodevelopmental variables* | 10 |
| Supplementary Figure 2 | *Questionnaire Data - Principal Components Analysis* | 11 |
| Supplementary Table 7 | *Extracted principal components with Eigenvalues >1* | 12 |
| Supplementary Table 8 | *Factor loadings from the rotated omnibus PCA matrix* | 13 |
| Supplementary Table 9 | *Inferential statistics for the K-prototype cluster solution* | 14 |
| Supplementary Figure 3 | *Distribution of gene functional networks and genetic diagnoses across the cohort-wide K-prototype cluster solution* | 15 |
| Supplementary Figure 4 | *Distribution of SVC genetic diagnoses and SVC sub-processes across the cohort-wide K-prototype cluster solution* | 16 |

### Supplementary Table 1

*Genetic diagnoses in the SVC group*

| Gene | N | % | Subprocess |
| --- | --- | --- | --- |
| STXBP1 | 26 | 23.85 | Docking and Priming |
| TRIO | 12 | 11.01 | Clustering and Scaffolding |
| SYT1 | 22 | 20.18 | Fusion |
| ATP6V1B2 | 1 | 0.92 | Fusion |
| KIF1A | 1 | 0.92 | Trafficking |
| NRXN1 | 2 | 1.83 | Clustering and Scaffolding |
| PRRT2 | 3 | 2.75 | Fusion |
| CACNA1A | 5 | 4.59 | Fusion |
| CLTC | 7 | 6.42 | Endocytosis |
| DNM1 | 2 | 1.83 | Endocytosis |
| SNAP25  CASK  RAB11B  SYP | 2  23  2  1 | 1.83  21.10  1.83  0.92 | Fusion  Clustering and Scaffolding  Endocytosis  Endocytosis |
| Total | 109 | 100 |  |

### Supplementary Table 2

*Genetic diagnoses in the non-SVC comparison group*

| Gene | N | % | FNG |
| --- | --- | --- | --- |
| ARID1B | 17 | 18.89 | Chromatin |
| CTNNB1 | 1 | 1.11 | Wnt signaling |
| DDX3X | 28 | 31.11 | Wnt signaling |
| DLG3 | 2 | 2.22 | Post-synaptic |
| DYRK1A | 2 | 2.22 | Post-synaptic |
| EHMT1 | 10 | 11.11 | Chromatin |
| KAT6B | 2 | 2.22 | Chromatin |
| PAK3 | 1 | 1.11 | Post-synaptic |
| SETD5 | 12 | 13.33 | Chromatin |
| SHANK3 | 3 | 3.33 | Post-synaptic |
| SMARCA2  ZDHHC9  GRIN2A  SYNGAP  GNAO1 | 8  1  1  1  1 | 8.89  1.11  1.11  1.11  1.11 | Chromatin  Post-synaptic  Post-synaptic  Post-synaptic  Post-synaptic |
| Total | 90 | 100 |  |

### Supplementary Table 3

*Demographic information and questionnaire descriptive data^a^*

|  | SVC | | | non-SVC group | | |
| --- | --- | --- | --- | --- | --- | --- |
|  | N | Mean (SD) | Range | N | Mean (SD) | Range |
| Age | 109 | 10.63 (6.43) | 2.67-32.33 | 90 | 12.92 (5.65) | 2.69-26.60 |
| Biological Sex (n) |  | Females 65 | Males 44 |  | Females 49 | Males 41 |
| Variant type (n) |  | PTV^b^ 34 | Missense 53 |  | PTV 16 | Missense 24 |
| VABS^29,30^ |  |  |  |  |  |  |
| Composite  Communication  Daily living  Socialisation  Motor | 88  97  97  97  84 | 48.53 (18.62)  42.72 (21.69)  47.20 (18.54)  50.18 (20.96)  51.82 (23.95) | 20-102  20-102  20-102  20-101  20-105 | 76  87  87  87  67 | 57.03 (17.58)  55.75 (21.95)  53.90 (18.97)  59.89 (18.80)  65.29 (16.46) | 20-118  20-107  20-127  20-110  20-121 |
| SRS^31^ |  |  |  |  |  |  |
| Total | 78 | 75.41 (11.75) | 43-112 | 71 | 76.37 (11.93) | 46-98 |
| Awareness | 85 | 73.80 (12.06) | 45-108 | 81 | 74.73 (11.44) | 50-98 |
| Cognition  Communication | 85  85 | 75.08 (10.64)  74.94 (11.94) | 39-106  40-106 | 81  81 | 72.49 (10.66)  74.41 (11.60) | 41-92  42-96 |
| Motivation  Restrictive, repetitive behaviour | 85  85 | 62.78 (12.63)  74.66 (14.55) | 41-104  43-108 | 81  81 | 67.25 (12.74)  78.62 (14.38) | 40-97  48-108 |
| DBC^32^ |  |  |  |  |  |  |
| Total | 85 | 55.12 (9.86) | 39-82 | 69 | 55.76 (13.38) | 34-106 |
| Disruptive | 92 | 51.82 (11.08) | 36-85 | 79 | 51.56 (13.56) | 35-112 |
| Self-absorbed  Communication  Anxiety  Social relating | 89  90  91  92 | 58.45 (11.11)  53.27 (13.12)  51.88 (10.04)  52.99 (10.15) | 38-93  36-100  36-80  38-84 | 78  79  79  79 | 59.36 (13.97)  58.63 (17.17)  52.86 (11.70)  52.89 (11.15) | 37-104  36-132  36-85  36-84 |
| CVI^33^ |  |  |  |  |  |  |
| Total | 86 | 15.46 (8.58) | 0-36 | 51 | 12.29 (6.86) | 0-32 |
| Visual attitude | 91 | 7.25 (4.87) | 0-19 | 56 | 5.55 (4.42) | 0-18 |
| Ventral stream  Dorsal stream  Complex problems  Other senses  Associated characteristics | 91  91  91  91  91 | 1.02 (1.20)  2.65 (2.50)  1.01 (.77)  1.15 (1.01)  2.41 (1.32) | 0-5  0-10  0-2  0-3  0-5 | 56  56  56  56  56 | .50 (.81)  1.88 (1.76)  1.16 (.78)  1.02 (1.02)  2.16 (1.55) | 0-4  0-6  0-2  0-3  0-6 |

^a^raw data prior to imputation; ^b^PTV = protein-truncating variant

| Supplementary Table 4  *Clinical phenotypes summary across the SVC-disorder group* | | | | |
| --- | --- | --- | --- | --- |
| Clinical feature  HPO Term Identifier^a^ | Data  available (n) | Frequency of Feature (n) | Frequency of Feature (%) | Subtype (n) |
| Delayed speech and language development  HP: 0000750 | 105 | 105 | 100 | Mild = using words and phrases (41); moderate = using single words only (12); severe = not using any words (41); unable to classify under age 5 years or insufficient information (11) |
| Abnormal eye physiology  HP: 0012373 | 103 | 61 | 59 | Strabismus/esotropia (15); nystagmus (4); hypermetropia (7); cerebral visual impairment (8); visual impairment unspecified (30) |
| Motor delay  HP: 0001270 | 105 | 85 | 81 | Mild = walked after 18 months and by 3 years (33); moderate = walked by 5 years (13); severe = walked after 5 years or nonambulatory over the age of 5 (25); unable to classify because nonambulatory under the age of 5 years (9); walking but age unspecified (4) |
| Abnormal muscle tone  HP: 0003808 | 104 | 75 | 72 | Hypotonia (64); hypertonia (19); tone unspecified (4) |
| Abnormality of movement  HP: 0100022 | 88 | 78 | 89 | Dystonia (20); chorea (21); dyskinesia (1); ataxia (64); tremor (24); stereotypies (40) |
| Sleep disturbance  HP: 0002360 | 106 | 73 | 69 | Hypersomnia (10); insomnia (13); frequent wakings (18); sleep disturbance unspecified (34) |
| Abdominal symptom  HP: 0011458 | 105 | 85 | 81 | Referring to feeding difficulties only |
| Self-injurious behaviour  HP: 0100716 | 106 | 63 | 59 | Finger biting or chewing (21); head banging (10); skin picking (7); other or unspecified (30) |
| Seizure  HP: 0001250 | 106 | 53 | 50 | Absence seizures (15); tonic-clonic seizures (19); infantile spasms (6); other or unspecified (23)    Seizure onset in infancy^b^ (28); seizure onset during childhood^c^ (25); seizures during infancy continued to childhood (25) |
| Abnormality of prenatal development or birth  HP: 0001197 | 106 | 25 | 24 | Mild prematurity (12); neonatal resuscitation (5); SCBU/NICU (19) |
| ID severity as measured by Vineland using DSM-5 levels^29,30^ | 109 | - | - | Borderline (18); mild (18); moderate (25); severe (27); profound (9) |

*^a^Human Phenotype Ontology*

*^b^Infancy is defined as 0-12 months of age.*

*^c^Childhood is defined as 1-18 years of age.*

### Supplementary Table 5

| *Clinical phenotypes summary across the non-SVC disorders group* | | | | |
| --- | --- | --- | --- | --- |
| Clinical feature  HPO Term Identifier^a^ | Data  available (n) | Frequency of Feature (n) | Frequency of Feature (%) | Subtype (n) |
| Delayed speech and language development  HP: 0000750 | 81 | 81 | 100 | Mild = using words and phrases (43); moderate = using single words only (13); severe = not using any words (19); unable to classify under age 5 years or insufficient information (6) |
| Abnormal eye physiology  HP: 0012373 | 80 | 40 | 50 | Strabismus/esotropia (16); nystagmus (3); hypermetropia (5); cerebral visual impairment (2); visual impairment unspecified (14) |
| Motor delay  HP: 0001270 | 81 | 71 | 88 | Mild = walked after 18 months and by 3 years (38); moderate = walked by 5 years (13); severe = walked after 5 years or nonambulatory over the age of 5 (9); unable to classify because nonambulatory under the age of 5 years (0); walking but age unspecified (1) |
| Abnormal muscle tone  HP: 0003808 | 80 | 56 | 71 | Hypotonia (50); hypertonia (10); tone unspecified (2) |
| Abnormality of movement  HP: 0100022 | 33 | 23 | 70 | Dystonia (5); chorea (4); dyskinesia (0); ataxia (18); tremor (4); stereotypies (14) |
| Sleep disturbance  HP: 0002360 | 81 | 48 | 59 | Hypersomnia (5); insomnia (6); frequent wakings (17); sleep disturbance unspecified (21) |
| Abdominal symptom  HP: 0011458 | 79 | 66 | 84 | Referring to feeding difficulties only |
| Self-injurious behaviour  HP: 0100716 | 80 | 32 | 40 | Finger biting or chewing (12); head banging (5); skin picking (5); other or unspecified (11) |
| Seizure  HP: 0001250 | 81 | 23 | 28 | Absence seizures (6); tonic-clonic seizures (5); infantile spasms (2); other or unspecified (11)    Seizure onset in infancy^b^ (7); seizure onset during childhood^c^ (15); seizures during infancy continued to childhood (7) |
| Abnormality of prenatal development or birth  HP: 0001197 | 81 | 17 | 21 | Mild prematurity (6); neonatal resuscitation (1); SCBU/NICU (15) |
| ID severity as measured by VABS using DSM-5 levels^29,30^ | 87 | - | - | Borderline (17); mild (33); moderate (23); severe (10); profound (3) |

*^a^Human Phenotype Ontology*

*^b^Infancy defined as 0-12 months of age.*

*^c^Childhood is defined as 1-18 years of age.*

### Supplementary Table 6

| *Comparison of clinical characteristics between SVC and non-SVC groups* | | |
| --- | --- | --- |
| Clinical domain | Significance* (p) | Cramer’s V (V) |
| Eye physiology | p = .233 | V = .092 |
| Tone abnormalities | p = .625 | V = .037 |
| Sleep disturbance | p = .217 | V = .100 |
| Feeding difficulties | p = .580 | V = .044 |
| Prenatal development/ perinatal problems  Seizures  Self-injurious behaviour  Movement disorders | p = .726  p = .003  p = .034  p = .025 | V = .031  V = .218  V = .191  V = .227 |

** Fisher’s Exact tests p = .05*

### Supplementary Figure 1

*Data missingness prior to imputation for all neurodevelopmental variables*

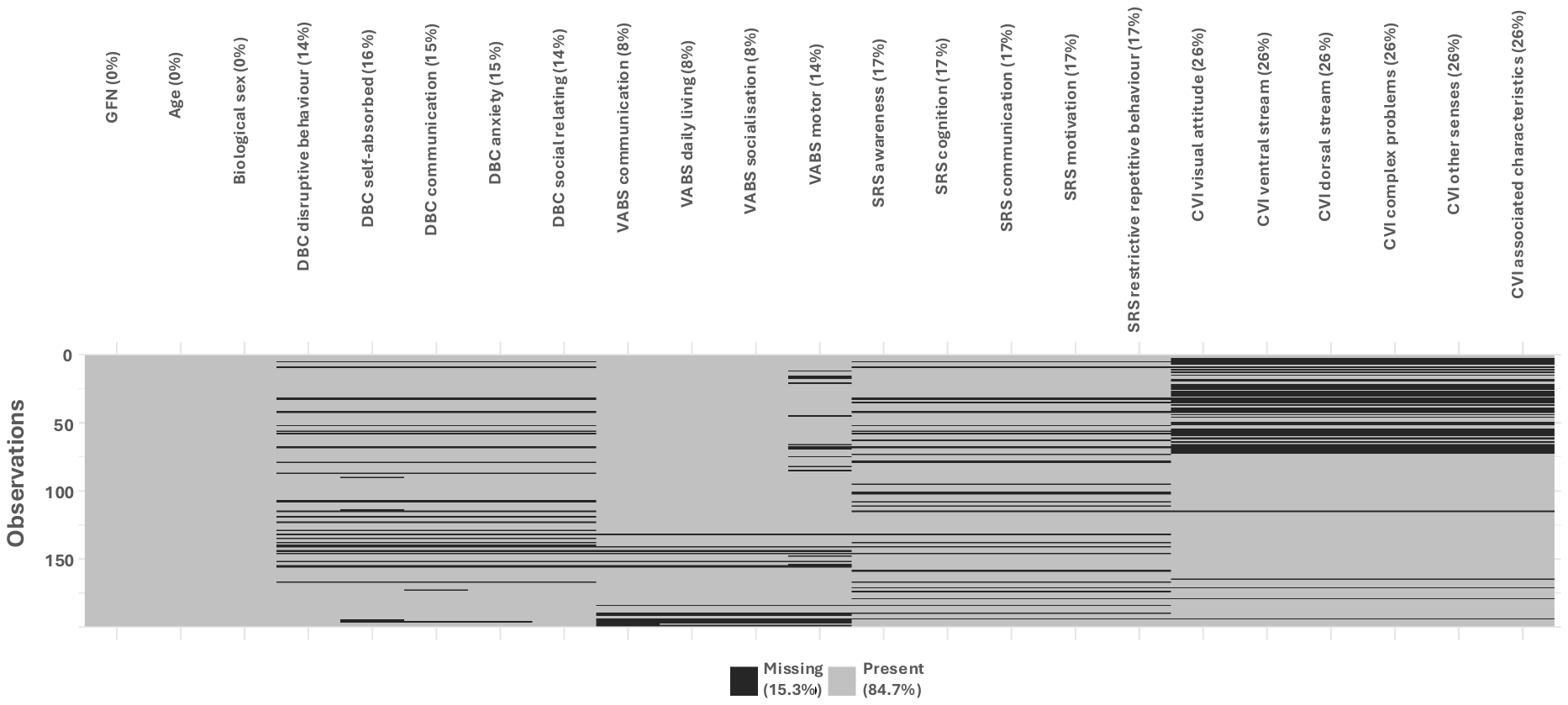
 Percentages in brackets on the top edge of figure represent missing data per variable.

### Supplementary Figure 2

*Questionnaire Data - Principal Components Analysis*

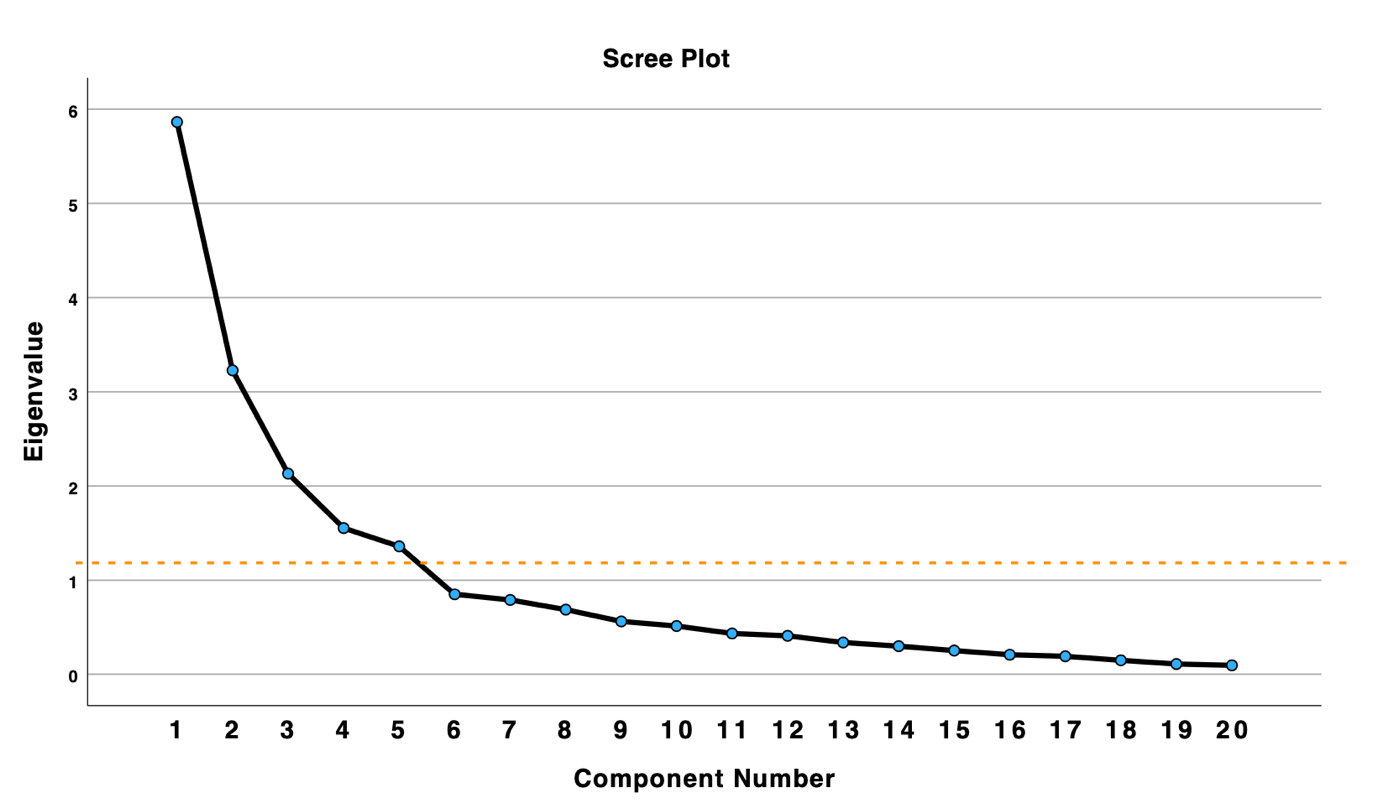

Scree plot, indicating an elbow and marked reduction in eigenvalues after five factors.

### Supplementary Table 7

| *Extracted principal components with Eigenvalues >1* | | | |
| --- | --- | --- | --- |
| Principal component | Eigenvalue | Explained variance (%) | Cumulative explained variance (%) |
| PC1 | 5.864 | 29.318 | 29.318 |
| PC2 | 3.227 | 16.137 | 45.455 |
| PC3 | 2.131 | 10.657 | 56.112 |
| PC4 | 1.553 | 7.767 | 63.879 |
| PC5 | 1.360 | 6.798 | 70.677 |

### Supplementary Table 8

| *Factor loadings from the rotated omnibus PCA matrix* | | | | | |
| --- | --- | --- | --- | --- | --- |
| Subdomain | Component | | | | |
|  | 1 | 2 | 3 | 4 | 5 |
| SRS Communication | **.903** | .156 | .077 | .120 | .058 |
| SRS Awareness | **.812** | .147 | .075 | .086 | -.017 |
| SRS Motivation | **.787** | -.102 | .119 | -.030 | .206 |
| SRS Cognition | **.770** | .245 | .245 | .077 | .102 |
| SRS RRB | **.768** | -.044 | .283 | .292 | .004 |
| DBC Socialisation | **.551** | .113 | .236 | -.018 | .452 |
| VABS Communication | .060 | **.924** | .055 | .095 | .149 |
| VABS Daily Living Skills | .081 | **.910** | .109 | .008 | .068 |
| VABS Socialisation | .168 | **.905** | .138 | .009 | .149 |
| VABS Motor skills | .057 | **.820** | -.095 | .248 | .120 |
| DBC Disruptive | .115 | .152 | **.891** | -.026 | -.012 |
| DBC Communication | .159 | .117 | **.820** | -.177 | -.134 |
| DBC Anxiety | .289 | -.143 | **.663** | .004 | .211 |
| DBC Self-absorbed | .230 | .061 | **.661** | .389 | .011 |
| CVI Other Senses | .044 | -.001 | .010 | **.739** | .283 |
| CVI Complex Problems | .120 | .109 | .080 | **.711** | -.169 |
| CVI Associated Characteristics | .105 | .107 | -.076 | **.644** | .092 |
| CVI Visual Attitude | .241 | .059 | .194 | -.066 | **.741** |
| CVI Ventral Stream | .033 | .253 | -.202 | .128 | **.726** |
| CVI Dorsal Stream | .031 | .236 | -.053 | .451 | **.630** |

Rotation Method: Varimax with Kaiser Normalisation.

Factor loadings exceeding .5 are marked in bold.

### Supplementary Table 9

| *Inferential statistics for the K-prototype cluster solution* | | |
| --- | --- | --- |
|  | Omnibus ANOVA/Chi-square | |
|  | Cohort-wide | SVC-only |
| **Demographic** |  |  |
| Age | F(3, 175) = 3.699, p = .013 | F(3, 96) = .338, p = .798 |
| Sex | x^2^ = 3.16, df = 3, p = .367, Cramer’s V = .133 | x^2^ = 1.377, df = 3, p = .711, Cramer’s V = .117 |
| Variant type | x^2^ = 4.55, df = 3, p = .208, Cramer’s V = .195 | X^2^ = 3.700, df = 3, p = .296, Cramer’s V = .215 |
| GFN distribution | x^2^ = 10.6, df = 3, p = .014, Cramer’s V = .244 | n/a |
| SVC-subprocess | n/a | x^2^ = 8.318, df = 12, p = .760, Cramer’s V = .760 |
| **Neurodevelopmental^a^** |  |  |
| Social-emotional | F(3, 175) = 2.401, p = .069 | F (3, 96) = 2.360, p = .076 |
| Adaptive functioning | F(3, 175) = 29.872, p = <.001 | F (3, 96) = 9.781, p = <.001 |
| Behavioural | F(3, 175) = 100.369, p = <.001 | F (3, 96) = 28.846, p = <.001 |
| Sensory-motor | F(3, 175) = 100.836, p = <.001 | F (3, 96) = 41.946, p = <.001 |
| Visual awareness | F(3, 175) = 82.908, p = <.001 | F (3, 96) = 35.590, p = <.001 |
| **Clinical** |  |  |
| Feeding difficulties | x^2^ = 15.893, df = 3, p = .001, Cramer’s V = .298 | x^2^ = 18.482, df = 3, p = <.001, Cramer’s V = .430 |
| Tone abnormalities | x^2^ = 6.09, df = 3, p = .107, Cramer’s V = .184 | x^2^ = 3.084, df = 3, p = .384, Cramer’s V = .175 |
| Sensory problems | x^2^ = 1.908, df = 3, p = .592, Cramer’s V = .103 | x^2^ = 6.228, df = 3, p = .101, Cramer’s V = .250 |
| Self-injury | x^2^ = 18.918, df = 3, p = <.001, Cramer’s V = .325 | x^2^ = 12.263, df = 3, p = .007, Cramer’s V = .350 |
| Epilepsy | x^2^ = 11.738, df = 3, p = .008, Cramer’s V = .256 | x^2^ = 5.826, df = 3, p = .120, Cramer’s V = .241 |
| Sleep difficulties | x^2^ = 19.754, df = 3, p = <.001, Cramer’s V = .332 | x^2^ = 11.206, df = 3, p = .011, Cramer’s V = .355 |
| Movement disorder | - | x^2^ = 5.117, df = 3, p = .163, Cramer’s V = .240 |

^a^P-values set at values ≤.01 across PCA-derived principal components (Bonferroni-corrected)

### Supplementary Figure 3

*Distribution of gene functional networks and genetic diagnoses across the cohort-wide K-prototype cluster solution*

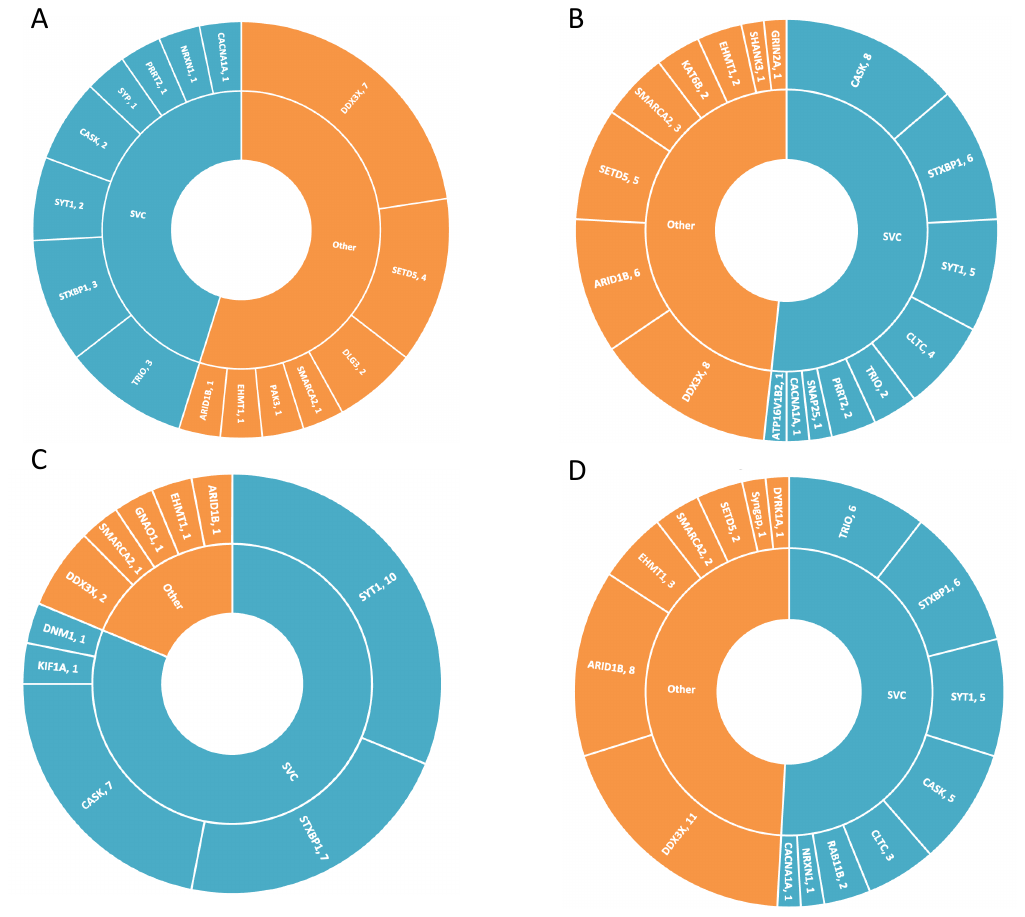

The SVC group is coloured in blue, and the non-SVC group is coloured in orange. (A) Cluster 1; (B) Cluster 2; (C) Cluster 3; (D) Cluster 4.

### Supplementary Figure 4

*Distribution of SVC genetic diagnoses and SVC sub-processes across the cohort-wide K-prototype cluster solution*

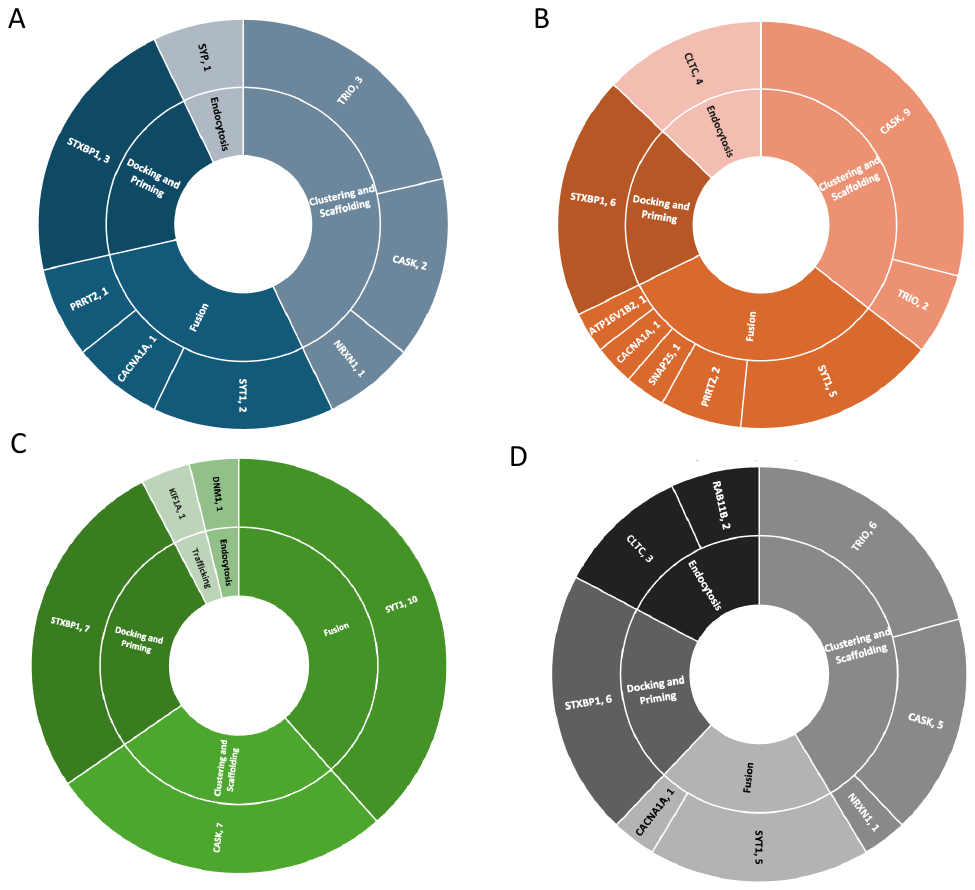

(A) Cluster 1; (B) Cluster 2; (C) Cluster 3; (D) Cluster 4.
